# High levels of serum cholesterol positively correlate with the risk of the development of vessel co-opting tumours in colorectal cancer liver metastases

**DOI:** 10.1101/2022.10.16.22281153

**Authors:** Miran Rada, Lucyna Krzywon, Audrey Kapelanski-Lamoureux, Stephanie Petrillo, Andrew R. Reynolds, Anthoula Lazaris, Nabil Seidah, Peter Metrakos

**Author notes:** Correspondence: Miran Rada. Cancer Research Program, McGill University, Montreal, Quebec, H4A3JI, Canada., Peter Metrakos. Cancer Research Program, McGill University, Montreal, Quebec, H4A3JI, Canada. These authors contributed equally to this work (Lucyna Krzywon, Audrey Kapelanski-Lamoureux).

## Abstract

Colorectal cancer liver metastatic (CRCLM) tumours present as two main histopathological growth patterns (HGPs) including desmoplastic HGP (DHGP) and replacement HGP (RHGP). The DHGP tumours obtain their blood supply by sprouting angiogenesis, whereas the RHGP tumours utilize an alternative vascularisation known as vessel co-option. In vessel co-option, the cancer cells hijack the mature sinusoidal vessels to obtain blood supply. Vessel co-option has been reported as an acquired mechanism of resistance to anti-angiogenic treatment in CRCLM. Here, we show the connection between the concentration of serum cholesterol and the development of vessel co-option in CRCLM. Our clinical data suggested that the elevation of serum cholesterol levels correlates with the risk of developing vessel co-opting tumours. Moreover, inhibition of the key modulators of cholesterol metabolism including HMGCR or PCSK9 attenuated the development of CRCLM tumours, as well as vessel co-option in vivo. Altogether, our data uncovered the importance of cholesterol in the development of vessel co-option tumours and demonstrated PCSK9 and HMGCR inhibitors as promising strategies to mitigate the development of vessel co-option tumours in CRCLM.

## Introduction

Colorectal cancer (CRC) is the third most common cancer and the second deadliest cancer^1^, which accounts for approximately 10% of cancer-related death in men and women^2^. The liver is the most common site of metastases of colorectal cancer, and approximately 50% of patients diagnosed with CRC develop liver metastases (LM) during the course of their disease^3^. Surgical resection improves survival by up to 50% and could be a potentially curative treatment for CRCLM patients^4^. However, only 15-20% of CRCLM patients are eligible for surgery resection^5^. The rest of the patients are treated with chemotherapy and targeted therapies such as anti-angiogenic agents (e.g. Bevacizumab)^6,7^. However, acquired resistance against these treatments frequently occurs that resulting in treatment failure and cancer recurrence^8^.

Colorectal cancer liver metastases (CRCLMs) present with two common histopathological growth patterns (HGPs): Desmoplastic HGP (DHGP) and replacement HGP (RHGP)^8–11^. The fundamental histopathological feature of DHGP lesions is the presence of a desmoplastic rim at the tumour periphery that separates cancer cells from liver parenchyma^8^. However, the desmoplastic rim is absent in RHGP lesions, and the cancer cells infiltrate through adjacent liver plates^8,12,13^. Moreover, angiogenesis is the predominant vascularization in DHGP tumours^8^. However, vessel co-option is the prevalent vascularization in RHGP tumours, in which the cancer cells co-opt the pre-existing vessels to obtain the blood supply^8,14^. Previous investigations reported vessel co-option as acquired resistance to anti-angiogenic therapy in various cancers including CRCLM, hepatocellular carcinoma (HCC) and glioblastoma^15^.

Various studies indicated a positive association between the risk of developing cancer and the elevation of serum cholesterol levels^16^. Cholesterol is an important lipid for supporting cellular homeostasis^17^. Cholesterol homeostasis maintenance depends on its biosynthesis and food intake^17,18^. Liver is the main organ for cholesterol biosynthesis^18^. Cholesterol is synthesized from acetyl-coenzyme A (CoA) through 30 enzymatic reactions, and the amount and activity of 3-hydroxy-3-methylglutaryl coenzyme A reductase (HMGCR) determine the levels of cholesterol synthesis^16^. Currently, different types of HMGCR inhibitors are approved as the first-line therapeutic drugs for lowering cholesterol levels, such as atorvastatin, rosuvastatin, pitavastatin, simvastatin, and pravastatin^19^. Previous investigations suggested repurposing of statins as anticancer agents against various cancers^20^. In this context, atorvastatin and simvastatin have been reported as an inhibitor of proliferation and inducer of apoptosis in breast cancer cells^21^. Additionally, it has been demonstrated that simvastatin increases the efficacy of chemotherapy and anti-angiogenic therapy in metastatic colorectal cancer patients^22^. However, the role of statins in the development of vessel co-option has not yet been determined.

Low-density lipoprotein (LDL) is the primary carrier of cholesterol in the blood to deliver cholesterol to peripheral and liver cells, and LDL receptor (LDLR) is the major determinant of LDL uptake by the liver^23^. The levels of LDL in the blood are controlled by proprotein convertase subtilisin-like kexin type 9 (PCSK9), which targets LDLR for lysosomal degradation^18,24^. Evolocumab is the monoclonal antibody against PCSK9 that binds to the circulating PCSK9 protein, inhibiting it from binding to the LDLR^25^. Evolocumab has been approved for the treatment of hypercholesterolaemia^26^. Recently, various preclinical studies suggested evolocumab treatment to increase the efficacy of anti-cancer agents, such as doxorubicin and trastuzumab^27^. Moreover, Liu et al.^28^ have reported that evolocumab potentiated immune checkpoint therapy against various types of tumours in vivo. However, the role of PCSK9 and its inhibitors in the development of vessel co-option is unknown.

In this manuscript, our clinical data suggested a positive correlation between serum levels of cholesterol and the development of vessel co-opting tumours in CRCLM. Our preclinical data also suggested atorvastatin and evolocumab as promising agents to attenuate the development of vessel co-option CRCLM tumours.

## Materials and methods

### Clinical data and patient samples

The study was conducted in accordance with the guidelines approved by McGill University Health Centre Institutional Review Board (IRB). Informed consent was obtained from all patients through the McGill University Health Centre (MUHC) Liver Disease Biobank. This study was performed on 84 CRCLM patients who were referred to McGill University Health Centre (MUHC) in Montreal. For patient characteristics see Table 1. Surgical specimens were procured and released to the Biobank immediately after the pathologist’s confirmation of carcinoma and surgical margins.

**Table 1.** Demographic baseline of CRCLM patients.

|  | Characteristic | Value | Percentage (%) |
| --- | --- | --- | --- |
| 1 | Sex |  |  |
|  | Male | 55 | 66.27 |
|  | Female | 29 | 33.73 |
| 2 | Age |  |  |
|  | <50 years | 4 | 4.82 |
|  | 50–70 years | 49 | 59.04 |
|  | >70 years | 30 | 36.14 |
| 3 | Body mass index category (BMI) |  |  |
|  | Underweight | 1 | 1.20 |
|  | Normal | 33 | 39.76 |
|  | Overweight | 26 | 31.33 |
|  | Obese | 21 | 25.30 |
|  | Insufficient data | 2 | 2.41 |
| 4 | Histopathological growth pattern (HGP)<br>(50% predominant HGP cut-off) |  |  |
|  | Replacement HGP (RHGP) | 35 | 42.17 |
|  | Desmoplastic HGP (DHGP) | 30 | 36.14 |
|  | Mixed | 18 | 21.69 |
| 5 | Histopathological growth pattern (HGP)<br>(100% predominant HGP cut-off) |  |  |
|  | Replacement HGP (RHGP) | 17 | 20.24 |
|  | Desmoplastic HGP (DHGP) | 11 | 13.10 |
| 6 | Neoadjuvant before liver resection |  |  |
|  | Yes | 66 | 78.57 |
|  | No | 16 | 19.05 |
|  | Insufficient data | 2 | 2.38 |
| 7 | Liver metastasis recurrence |  |  |
|  | Yes | 27 | 32.14 |
|  | No | 42 | 50.00 |
|  | Never fully resected | 15 | 17.85 |
| 8 | Hypocholesterolemic agent at time of surgery |  |  |
|  | Yes | 23 | 27.38 |
|  | No | 61 | 72.62 |
| 9 | Type of hypocholesterolemic agent |  |  |
|  | Statin | 21 | 25.00 |
|  | Ezetimibe | 2 | 2.38 |

### Kaplan-Meier estimates of overall survival

Overall survival estimates were calculated from the date of diagnosis of liver metastases to the date of death or to the date of the last follow-up.

### Cell culture

Mouse colorectal cancer (MC38) cells were cultured in DMEM (Wisent Inc., #319-005-CL) supplemented with 10% FBS (Wisent Inc., #085-150) and 1× penicillin/streptomycin (Wisent Inc., 450-201-EL). All cells were cultured at 37 °C with 5% CO2.

### Immunohistochemical Staining

We performed immunohistochemical staining for formalin-fixed paraffin-embedded (FFPE) specimens as described in previous publications^9,12^. We performed staining using the following antibodies: HMGCR 1:100 (Thermo Fisher, #13533-1-AP), in-house polyclonal PCSK9 antibody^29^ 1:1000 (was a generous gift from Dr Nabil Seideh).

### Immunofluorescence staining

Immunofluorescence staining was performed for (FFPE) human CRCLM specimens as described in previous publications^30,31^. We used anti-LDLR antibody 1:200 (R&D systems, AF2255).

### Xenograft experiments

CRCLM tumours were generated in C57B/6 mice using intrasplenic injection protocol as described in previous publications^31,32^. For atorvastatin experiment, the mice were divided into 4 groups: A. Control (vehicle only), B. Treated for two weeks (40mg/kg every day) before intrasplenic injection. C. Treated for two weeks (40mg/kg every day) after intrasplenic injection. D. Treated (40mg/kg every day) for two weeks before intrasplenic injection and two weeks after intrasplenic injection.

For the PCSK9 inhibition experiment, the mice were divided into two groups after intrasplenic injection. The mice in the first group were injected with a vehicle alone (Control). The mice in the second group were treated with 200μg of evolocumab (also called Repatha; Amgen Manufacturing Limited) every two days. The evolocumab was a generous gift from Dr Nabil Seideh. The treatment started 5 days after intrasplenic injection.

### Statistical reproducibility

Statistical analysis was performed using GraphPad Prism software version 7.0 (GraphPad Software, La Jolla, CA, USA) software. Unpaired Student’s t-test was applied to compare the means of two groups. The association between the two categorical groups was assessed with the Chi-square test. For the overall survival data, the Log-Rank test was used to determine the statistical significance. P-values of <0.05 were considered to be significant. Data presented as mean ± standard deviation.

## Results

### Preoperative serum total cholesterol and LDL score as a novel predictor of the type of HGPs and overall survival in CRCLM patients

To investigate the association between serum cholesterol levels in CRCLM patients and the presence of vessel co-option tumours, we analyzed the total cholesterol and LDL in 84 CRCLM patients, of whom 66.27% were male and 33.73% were female (Table 1). Based on the 50% cut-off predominant HGPs scoring of the tumours, the patients were divided into three groups: predominant vessel co-opting RHGP (42.17%), predominant angiogenic DHGP (36.14%), and mixed (21.69%). The patients in the mixed group had both vessel co-opting and angiogenic tumours.

As shown in Figure 1a, the total cholesterol was significantly higher in patients of the vessel co-opting RHGP group compared to their angiogenic and mixed counterparts. We further analyzed the cohort and investigated the levels of total cholesterol in the patients that had 100% predominant vessel co-opting RHGP (n=17) or angiogenic DHGP (n=11) tumours (Figure 1b). Again, our results confirmed that the patients with RHGP tumours have significantly higher levels of total cholesterol in their serum than the patients with angiogenic tumours. Similar results were demonstrated for LDL (Figure 1c and d).

**Figure 1.**
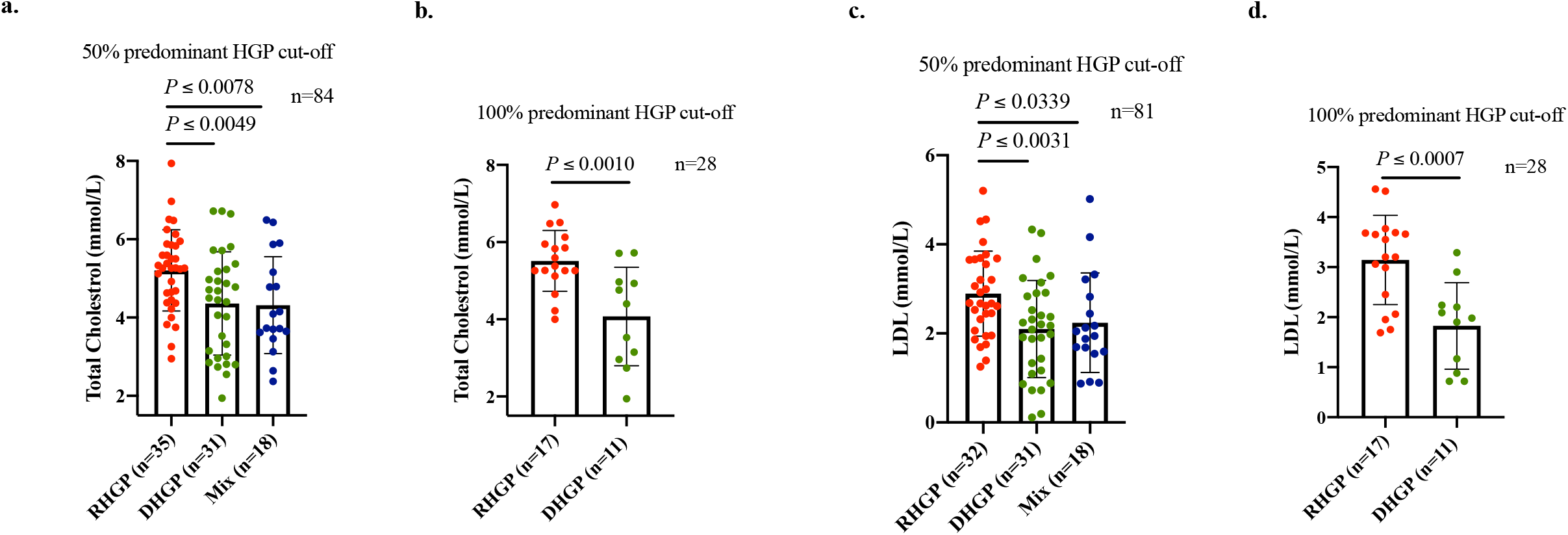
The levels of serum total cholesterol and LDL in CRCLM patients before surgical resection. **a**. Represents the levels of serum total cholesterol in patients with RHGP (n=35), DHGP (n=31) or mixed (n=18) tumours. The HGPs of the tumours were scored based on a 50% predominant HGP cut-off. **b**. Represents the levels of serum total cholesterol in patients with RHGP (n=17) or DHGP (n=11) CRCLM tumours. The HGPs of the tumours were scored based on a 100% predominant HGP cut-off. **c**. Shows the LDL levels in the serum of patients with RHGP RHGP (n=32), DHGP (n=31) or mixed (n=18) tumours. The HGPs of the tumours were scored based on a 50% predominant HGP cut-off. **b**. Shows the concentration of serum LDL in patients with RHGP (n=17) or DHGP (n=11) tumours. The HGPs of the tumours represent a 100% predominant HGP cut-off. Data are presented as the mean ± SD.

Next, we categorized the patients into four groups based on the levels of their total cholesterol (< 2.99, 3.00-3.99, 4.00-4.99, > 5.00) and LDL (< 0.99, 1.00-1.99, 2.00-2.99, > 3.00) (Figure 2). Interestingly, we observed a significant increase in the proportion of patients with vessel co-opting tumours upon the elevation of the total cholesterol or LDL levels (Figure 2a and b). Importantly, the patients with lower levels of total cholesterol or LDL had higher 5-year overall survival rates than those who had higher levels of total cholesterol or LDL (Figure 2c and d).

**Figure 2.**
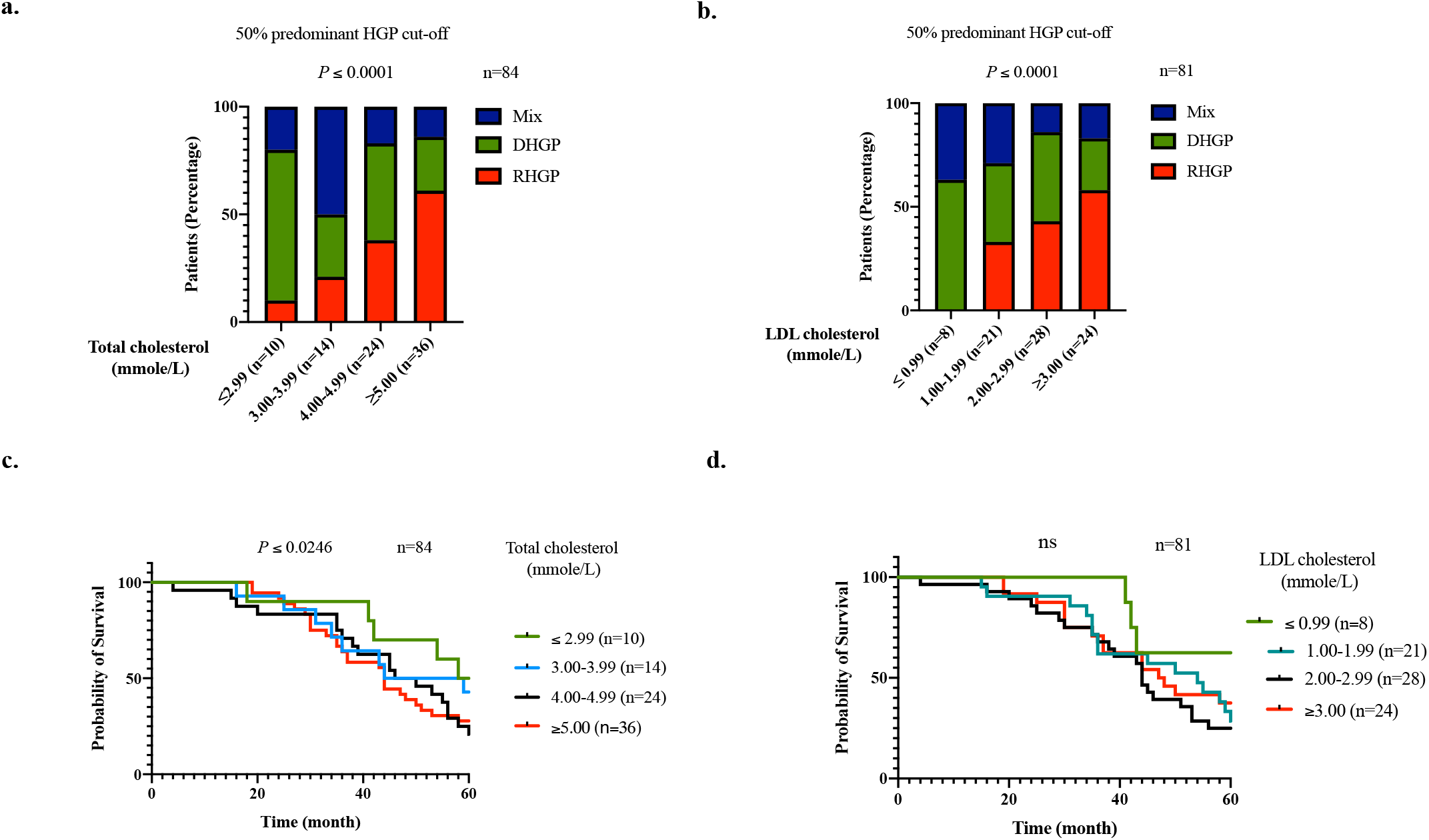
The concentration of serum total cholesterol and LDL positively correlates with the presence of vessel co-opting tumours in CRCLM patients. **a. and b**. The CRCLM patients were divided into different groups based on the concentration of serum total cholesterol and LDL, respectively. Each bar represents the percentage of CRCLM patients with different types of HGPs. **c. and d**. Kaplan-Meier estimates of 5-year overall survival for CRCLM patients according to serum total cholesterol (n=84) and LDL (n=81) levels. The HGPs of the tumours were scored based on a 50% predominant HGP cut-off.

Based on National Cholesterol Education Program (NCEP) guidelines, a serum total cholesterol concentration above 5.2 mmole/L is considered as high and undesirable^33^. Therefore, we used this guideline to further corroborate the association between the levels of serum total cholesterol and HGP of CRCLM tumours. Subsequently, we analyzed the type of HGPs (RHGP, DHGP or mixed) of the CRCLM tumours in the patients that had normal levels (< 5.2 mmole/L) or high levels (> 5.2 mmole/L) of serum total cholesterol. As shown in Figure 3a, the proportion of patients with vessel co-opting RHGP tumours was significantly higher in the group that had undesirable levels of serum total cholesterol compared to those patients that had lower levels of serum total cholesterol. Intriguingly, the 5-year overall survival of the patients that had a normal range of serum total cholesterol was significantly higher than their counterparts (Figure 3b). The results were more prominent when we increased the score of the predominant HGPs cut-off from 50% to 100% (Figure 3c and d).

**Figure 3.**
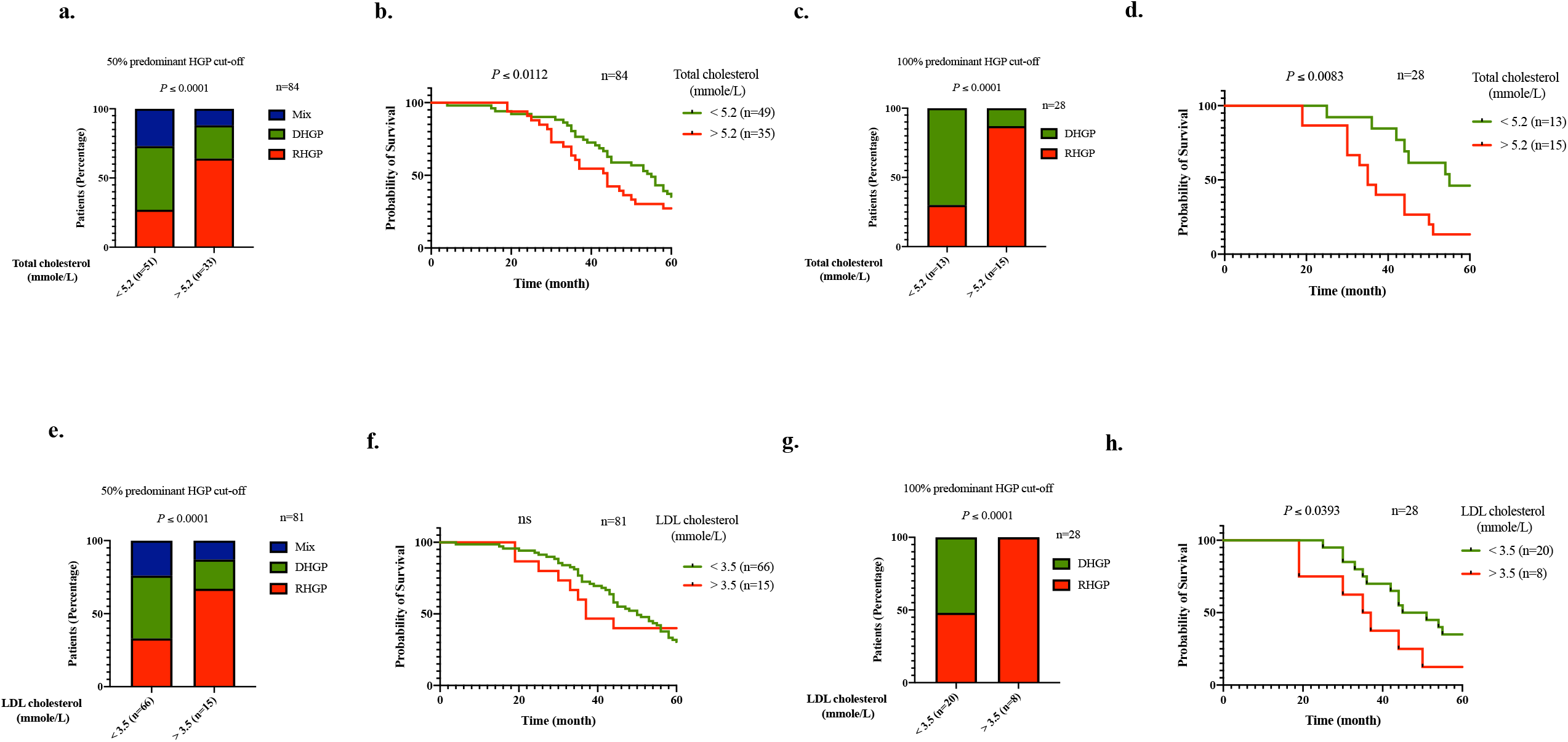
Preoperative serum total cholesterol and LDL score as a novel predictor of the type of HGPs and survival in CRCLM patients. **a and e**. Graphs show the percentage of patients with vessel co-option (RHGP), angiogenic (DHGP) or % mixed (RHGP and DHGP) lesions according to the concentration of serum total cholesterol or LDL, respectively. **b and f**. Kaplan– Meier survival curves showing 5-year overall survival in CRCLM patients according to serum total cholesterol or LDL levels, respectively. The HGPs of the tumours in the presented patients scored based on a 50% predominant HGP cut-off. **c and g**. Represent the percentage of patients with vessel co-option (RHGP) or angiogenic (DHGP) tumours according to the concentration of serum total cholesterol. **d and h**. Kaplan– Meier survival curves showing 5-year overall survival in CRCLM patients according to serum total cholesterol and LDL concentrations. The HGPs of the tumours were scored based on a 100% predominant HGP cut-off for patients.

Serum LDL-cholesterol (LDL) levels higher than 3.5 mmol/L are undesirable and classified as hypercholesterolemia^34^. Therefore, we used the cut-off of 3.5 mmol/L to classify the CRCLM patients into two groups including the patients with low LDL (n=66) and high LDL (n=15). We noticed an increase in the percentage of the patients with vessel co-opting tumours from 33% to 67% in response to increasing serum LDL (Figure 3e). Also, the 5-year overall survival was dramatically lower in the patients with higher levels of serum LDL (Figure 3f). Intriguingly, the association between LDL levels and the presence of vessel co-opting tumours was more significant when we repeated our analysis using the data of CRCLM patients with 100% predominant HGPs (Figure 3g and h). The percentage of CRCLM patients with RHGP lesions was 48% in low LDL (<3.5 mmol/L) group, while this percentage was increased to 100% in the group that had high levels (>3.5 mmol/L) of LDL (Figure 3g). Taken together, these data propose that high levels of serum total cholesterol and LDL are strongly associated with the presence of vessel co-opting RHGP tumours in CRCLM patients.

### Atorvastatin attenuates the development of vessel co-opting liver metastatic tumours

The primary source of cholesterol synthesis is the liver, and HMGCR serves as a key regulatory enzyme controlling this process^35^. Therefore, we decided to assess the expression levels of HMGCR in chemonaïve CRCLM specimens (RHGP, n=5; DHGP, n=5) by immunohistochemistry. Indeed, our data showed dramatic upregulation of HMGCR in the liver parenchyma of vessel co-opting RHGP sections compared to their angiogenic DHGP counterparts (Figure 4a). Statins are the inhibitor of HMGCR and have been used clinically to treat patients with high levels of cholesterol^36^. We noticed that some patients in our cohort also used statins during the course of their disease. Thus, we decided to categorize the patients into two groups including the patients who have not administrated statins (n=63) and those that used stains (n=21). Importantly, we demonstrated a significant reduction in the proportion of patients with vessel co-opting CRCLM lesions upon administration of statins (Figure 4b). Additionally, the patients who administrated statins had a significantly better survival rate compared to the rest of the patients (Figure 4c). To further examine the association between vessel co-option and statin administration in CRCLM patients, we compared the levels of total cholesterol or LDL in the patients treated with statins to the rest of the patients. The administration of statins significantly lowered the total cholesterol and LDL in the serum of patients with DHGP (n=10) and mixed lesions (n=5) (Figure 4d and e), while there was no significant difference between patients with RHGP (n=6) tumours and the rest of the patients (n=63). These results suggest that the administration of statins failed to achieve the reduction of total cholesterol and LDL in patients with RHGP tumours. We hypothesize that the failure of lowering cholesterol likely contributed to the development of vessel co-opting tumours. Further investigations are required to confirm this hypothesis.

**Figure 4.**
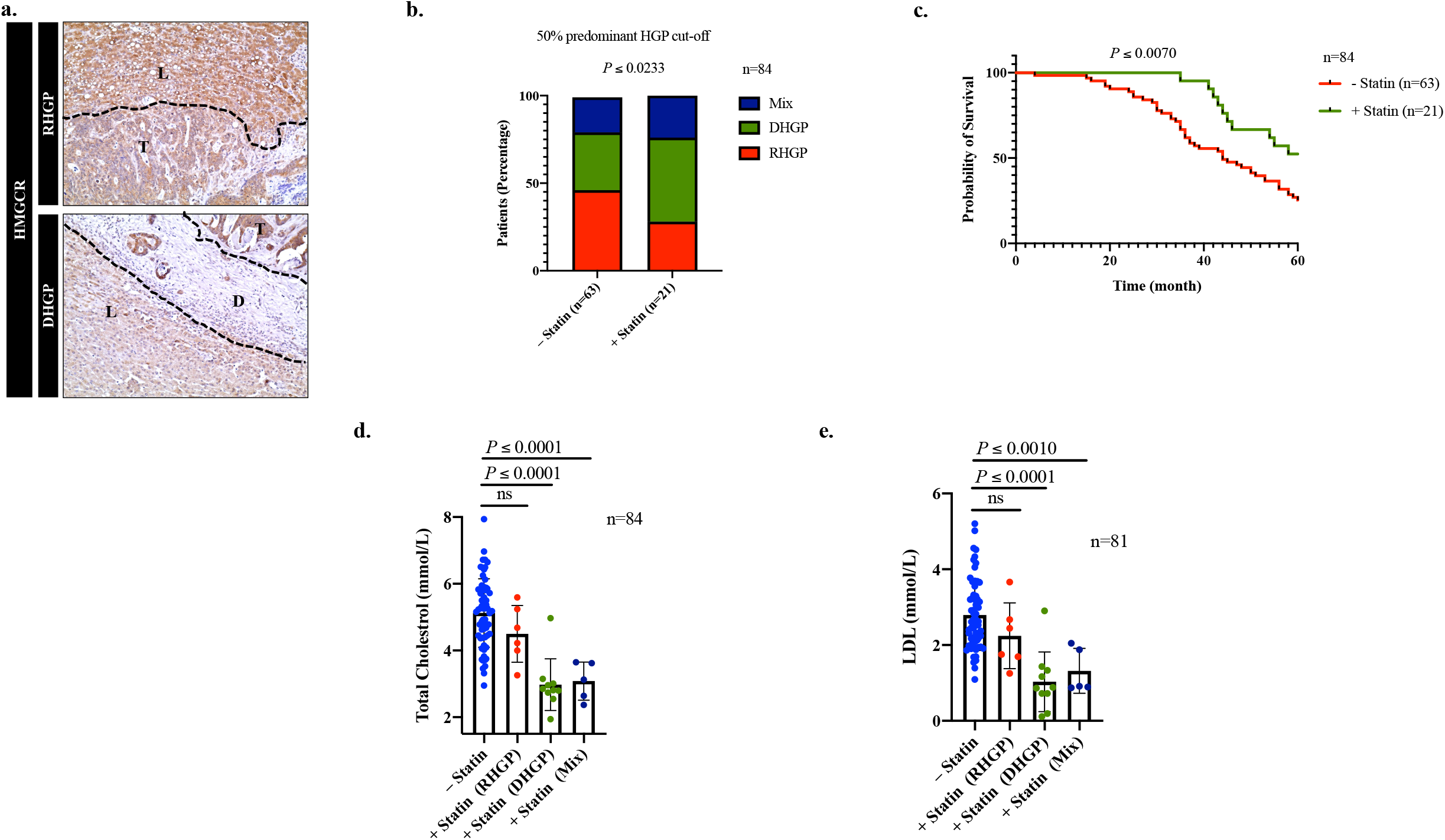
Inhibition of HMGCR associated with the presence of angiogenic tumours in CRCLM patients. **a**. Immunohistochemistry staining of CRCLM sections using an anti-HMGCR antibody. **b**. The percentage of CRCLM patients with various types of HGPs in the presence or absence of treatment with statins. **c**. Kaplan– Meier survival curves showing 5-year overall survival of CRCLM patients in the presence or absence of treatment with statins. **d and e**. Represent the levels of serum total cholesterol and LDL, respectively, of CRCLM patients in the presence or absence of treatment with statins. The treated patients with statins are divided into 3 groups according to the HGP of their tumours including RHGP (n=6), DHGP (n=10) and mixed (n=5). Data are presented as the mean ± SD.

Next, we performed in vivo experiments to validate our clinical data using atorvastatin. We generated CRCLM tumours by injecting MC38 colorectal cancer cells intrasplenically^31,37^. The mice were divided into four groups: A. control, B. treated with atorvastatin before intrasplenic injection, C. the mice were treated with atorvastatin after intrasplenic injection, D. the mice were treated with atorvastatin before and after intrasplenic injection. Generally, the treatment with atorvastatin weakened the generation, as well as the number and diameter of the generated tumours (Figure 5a). However, the best results were obtained with group D (Figure 5b and c), in which the mice were treated before and after intrasplenic injection. Later, we assessed the effect of atorvastatin treatment on the development of vessel co-option tumours by staining the harvested livers with hematoxylin and eosin (H&E). Accordingly, all generated tumours in control and group B were vessel co-option RHGP, while we demonstrated the development of the angiogenic DHGP tumours in the mice of C and D groups (Figure 5d and e). However, the strongest attenuation in the development of RHGP vessel co-option was found for the mice in group D. Altogether, our data suggested treatment with statins as a potential therapeutic strategy to reduce the development of vessel co-option CRCLM tumours.

**Figure 5.**
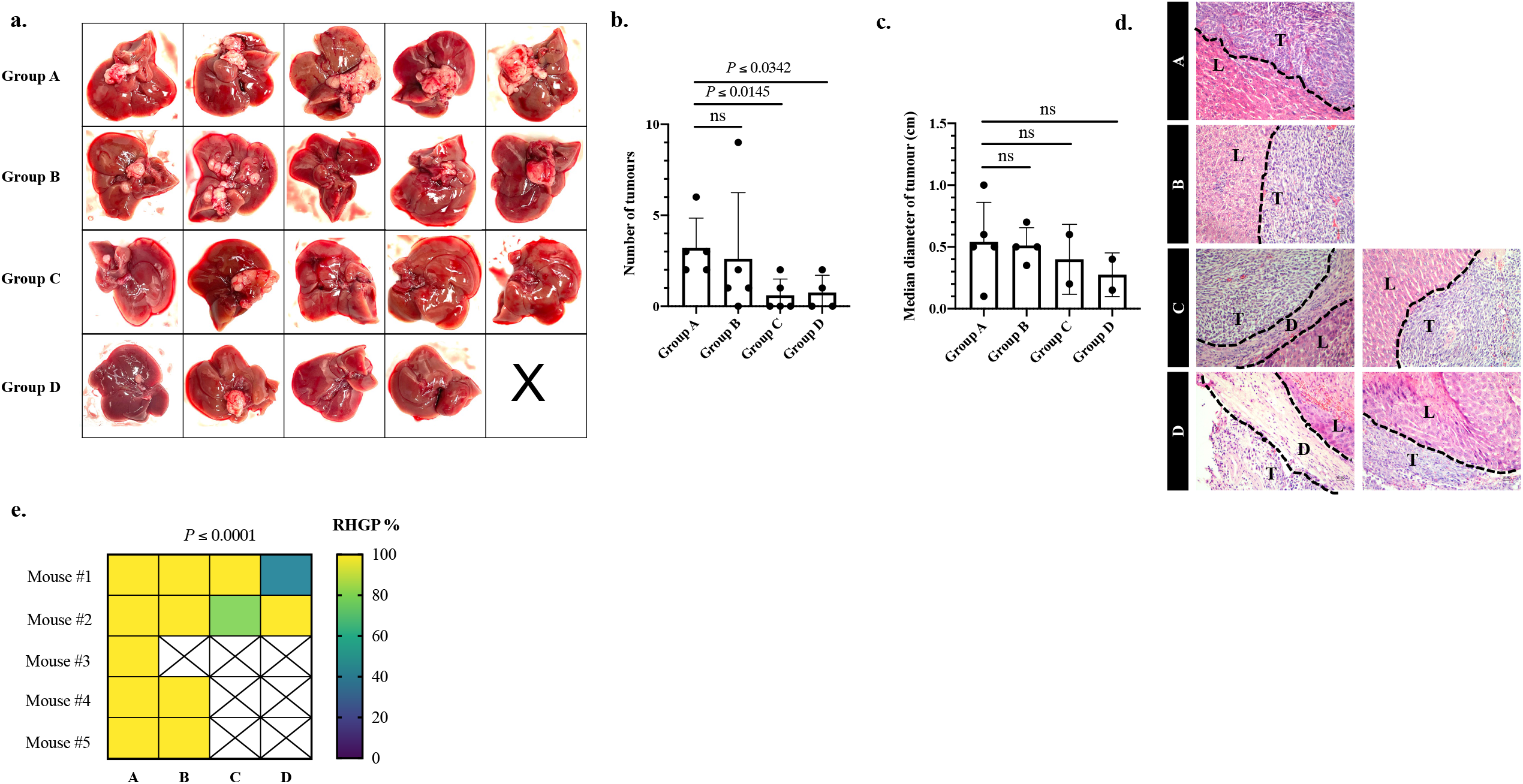
Treatment with atorvastatin deteriorates the development of vessel co-opting lesions in vivo. **a**. Images of the liver of the mice that received intrasplenic injections of MC-38 cells in the presence or absence of atorvastatin treatment. Group A: control (no treatment) mice, Group B: treated mice with atorvastatin for 2 weeks before intrasplenic injection of MC38 cells, Group C: treated mice with atorvastatin for 2 weeks after intrasplenic injection of MC38 cells, Group D: treated with atorvastatin for 2 weeks before and after intrasplenic injection of MC38 cells. **b** and **c**. Quantitation of the number and diameter of the generated liver metastatic tumours, respectively. **d**. H&E staining of metastatic lesions. T: tumour, D: desmoplastic ring, L: liver parenchyma. **e**. Shows the percentage of vessel co-opting RHGP in the generated tumours. Data are presented as the mean ± SD.

### Inhibition of PCSK9 via evolocumab reduces the establishment of vessel co-option tumours

Another mechanism that involved in hyperlipidemia is the overexpression of PCSK9, which degrade LDLR and reduce the absorption of LDL by liver^29,38^. Our immunohistochemical analysis showed a significant upregulation of PCSK9 in vessel co-opting RHGP specimens (n=5) compared to their angiogenic DHGP (n=5) counterparts (Figure 6a). Interestingly, the levels of LDLR were significantly reduced in the RHGP lesion (Figure 6b), which confirms the effect of PCSK9 upregulation. Therefore, we questioned whether PCSK9 contributes to the development of vessel co-opting CRCLM tumours. To answer our question, we intrasplenically injected C57B/6 mice with MC38 cells^31,37^ followed by treatment with evolocumab, an anti-PCSK9 monoclonal antibody that is approved to treat hyperlipidemia in human patients^25,39^. After intrasplenic injection with MC38, the mice were injected with either evolocumab (200μg/ every 2 days) or vehicle for two weeks. Significantly, treatment with evolocumab resulted in a significant reduction in the generation, number, and diameter of CRCLM tumours (Figure 6c-e). More importantly, the treatment significantly promoted the development of angiogenic DHGP lesions (Figure 6f and g). In sum, our data proposed PCSK9 as a promising therapeutic target to decrease the development of vessel co-opting CRCLM tumours.

**Figure 6.**
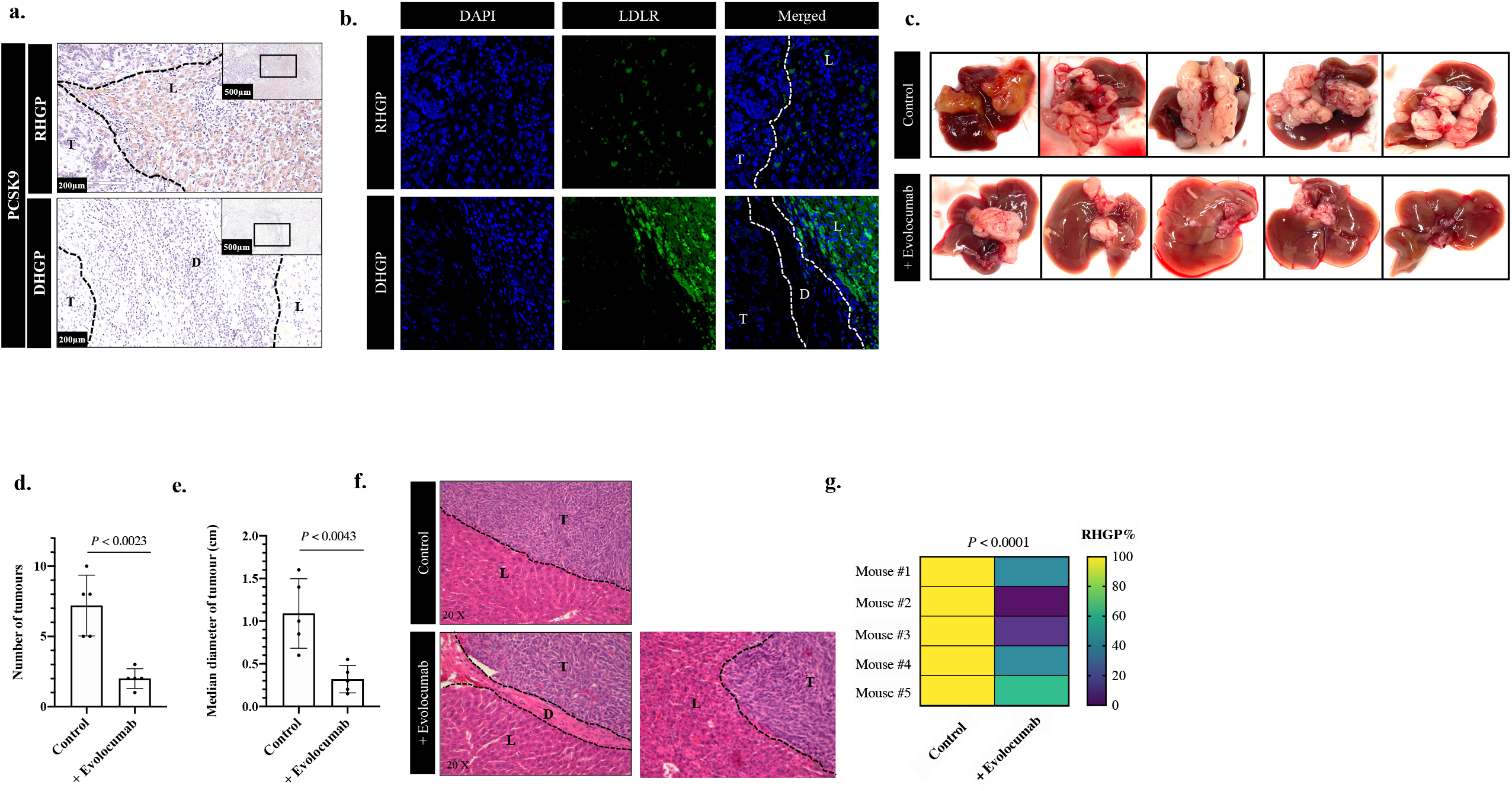
Inhibition of PCSK9 through evolocumab stimulates the development of angiogenic CRCLM tumours in vivo. **a**. Immunohistochemical staining of CRCLM sections using an anti-PCSK9 antibody. **b**. LDLR immunofluorescence staining of CRCLM sections. **c**. Images of the liver of control (untreated mice) and treated mice with anti-PCSK9 (Evolocumab) that received intra-splenic injections of MC-38 cells. **d** and **e**. Represent the number and diameter of the generated liver metastatic tumours, respectively. **f**. H&E staining of the generated lesions. **g**. Show the percentage of vessel co-option (RHGP) in the generated tumours upon the treatment with evolocumab.

## Discussion

Vessel co-option is a non-angiogenic vascularization method that allows the tumours to further their growth and spread by hijacking the pre-existing vasculature^11^. Therefore, the tumours that utilize vessel co-option vascularization are resistant to anti-angiogenic therapy^15^. Vessel co-option is a predominant vascularization in CRCLM^8,11,31^. We previously reported that CRCLM patients with predominantly angiogenic metastasis receiving neoadjuvant anti-angiogenic plus chemotherapy have significantly higher 5-year overall survival compared to the patients with co-opting tumours who have received the same treatment^8^. Recently, we found different molecules that facilitate the formation of vessel co-option, such as runt related transcription factor-1 (RUNX1)^31^, transforming growth factor beta-1 (TGFβ1)^10,31,37^, and Angiopoietin-1 (Ang1)^32^. However, the clinical benefit of inhibiting these molecules is not yet elucidated due to the lack of approved drugs against them in human patients. In this manuscript, we demonstrated a positive correlation between hyperlipidemia and vessel co-option in CRCLM, and our preclinical data suggested both HMGCR and PCSK9 as key molecules in this process. Interestingly, the inhibitors against both molecules are approved in human patients.

Elevated serum total cholesterol and LDL levels have been reported in various cancers, such as breast^40^, colon, gastric and ovarian^41^. Interestingly, Pelton et al.^42^ have suggested hypercholesterolemia promotes tumour angiogenesis in breast cancer. Similar results were proposed in hepatocellular carcinoma^43^ and prostate cancer^44^. In contrast, our results suggested that hypercholesterolemia impairs the development of angiogenic tumours and induces non-angiogenic vessel co-option in CRCLM. However, the molecular mechanisms by which hypercholesterolemia impairs angiogenesis in CRCLM are unknown. It is worth mentioning that hypercholesterolemia has been shown to inhibit angiogenesis via the inhibition of endothelium-derived nitric oxide (EDNO) release^38^, which is known as a mediator of angiogenesis^45^.

Our preclinical and clinical data showed that treatment with statin favours the development of angiogenic tumours in CRCLM. In agreement with our findings, Weis et al.^46^ observed proangiogenic effects of statin treatment in lung cancer, using in vitro and in vivo models. Similarly, Mascitelli et al.^47^ have shown demonstrated that simvastatin promotes VEGF production via RhoA downregulation and HIF-1α upregulation in endothelial cells to support tumour growth. In contrast, other studies reported the anti-angiogenic effect of statins in different types of tumours, such as glioblastoma^48^ and breast cancer^49^. These data suggest that reports about the role of statins in the inhibition of tumour angiogenesis are conflicting. It has been suggested that low concentrations of statin treatment have pro-angiogenic activities, while high concentrations have anti-angiogenic activities^50^. The effect of statins in angiogenesis is linked to endothelial cell proliferation, migration and apoptosis, as well as its involvement in vascular endothelial growth factor (VEGF) synthesis, the key angiogenic mediator^46,51–53^. Therefore, further investigations are required to confirm the effect of statins on vessel co-option in various cancers. The involvement of PCSK9 in tumour progression and resistance against anti-cancer agents has been investigated previously. In this context, overexpression of PCSK9 has been linked to poor prognosis in various cancers, such as HCC,^54^ CRC^55^, prostate cancer^56^, and ovarian cancer^57^. The function of PCSK9 in tumour angiogenesis is poorly understood. Abdelwahed et al.^56,58^ have reported that inhibition of PCSK9 suppressed tumour angiogenesis in prostate cancer. Interestingly, another study has linked PCSK9 overexpression to resistance against sorafenib, an anti-angiogenic agent, in HCC^59^. However, this study has not examined the role of PCSK9 in vessel co-option. Herein, we suggested a positive correlation between PCSK9 upregulation and vessel co-option in CRCLM. However, further studies are required to identify the role of PCSK9 in the development of vessel co-option in other types of cancers including glioblastoma and HCC.

## Conclusions

Our clinical data demonstrated that high levels of serum total cholesterol and LDL are associated with the risk of vessel co-option in CRCLM. Additionally, our data suggested HMGCR and PCSK9 as key molecules of hyperlipidemia in vessel co-opting CRCLM, and their inhibition attenuated the development of vessel co-option in vivo. To our knowledge, this is the first report to show the function of HMGCR and PCSK9 in the development of vessel co-option, specifically in CRCLM. These data have great translational potential. Firstly, our data suggest the possibility of using serum cholesterol data profiles to predict the presence of vessel co-option tumours. Also, our results demonstrate that targeting HMGCR and PCSK9 provides is a compelling rationale for conducting future clinical trials in CRCLM patients, particularly those patients that have resistant tumours to anti-angiogenic therapy.

## Data Availability

All data produced in the present study are available upon reasonable request to the authors

## Author Contributions

M.R., A.L., A.R., N.S. and P.M. co-conceived the study. L.K. and S.P. collected the clinical data. S.P. prepared CRCLM samples. M.R. analyzed the clinical data and performed cell culturing, immunohistochemistry, immunofluorescence, xenograft experiments, mice treatment, data curation, writing and preparation of the manuscript. A.K.L. assisted in the xenograft experiments. P.M. review the manuscript and funding acquisition.

## Institutional Review Board Statement

The study was approved by the McGill University Health Centre Institutional Review Board.

## Data Availability Statement

All data produced in the present study are available upon reasonable request to the authors

## Acknowledgments

The authors would like to acknowledge the support provided by Dana Massaro and Ken Verdoni Liver Metastases Research Fellowship.

